# Investigating the levels of polychlorinated biphenyls (PCBs) at three lactational stages: A comprehensive study in Ghana

**DOI:** 10.1101/2023.07.31.23293448

**Authors:** Justice Wiston Amstrong Jonathan, Daniel Elorm Kwame Kabotso, David K. Essumang, John K. Bentum, Francis Ofosu-Koranteng, Hintermann Mbroh, John Tampuori

**Affiliations:** Department of Chemistry, University of Cape, UCC; Department of Basic Sciences, School of Basic and Biomedical Sciences, UHAS; Pesticide Residues Laboratory, Ghana Standards Authority, GSA; Department of Gynaecology, Ho Teaching Hospital, HTH

**Keywords:** polychlorinated biphenyl, colostrum, transition milk, mature milk, congener, neonate

## Abstract

**Introduction:** The human breast secretes three types of milk during lactation – colostrum, transitional milk and mature milk However, like any other biofluid, human breast milk is not pristine

**Objectives:** The objectives of this study was to determine the levels of polychlorinated biphenyls congeners in colostrum, transitional milk and mature milk of mothers at the Ho Teaching Hospital to ascertain which portion contained the highest levels of PCPBs that could pose any significant health risks to neonates.

**Methodology:** A cross-sectional study design was employed to conduct the study at the Ho Teaching Hospital. Protocol for the study was reviewed and approved by the University of Health and Allied Sciences Research Ethics Committee (UHAS–REC). The study recruited forty-seven (47) mothers using purposive sampling. About 10.0 g of each aliquot of colostrum, transitional milk and mature milk were treated using a modified form of QuEChERS to extract PCBs and the resulting extract analyzed for fourteen (14) PCB congeners using GC–MS/MS.

**Results:** Fourteen (14) PCB congeners were examined in all the 141 breast samples, which represented three different types of human milk. All samples had levels that were below thresholds of detection. The lack of PCB residues in the examined samples may be viewed as positive news. It might suggest that the mothers were only minimally exposed to these industrial pollutants.

**Conclusion:** At any stage of lactation, no PCBs were found in the breast milk samples. As a result, there were no obvious health concerns for breastfed infants from the levels of PCBs found in the samples of breast milk that were examined. The result is good news for international efforts to eliminate PCBs in the environment and on people.

## Introduction

The importance of human breast milk as an ideal source of nutrition for the healthy growth and development of infants can never be underestimated. Accordingly, health organizations around the world continue to support and promote breastfeeding among mothers. It is in recognition of this fact that the World Health Organization has recommended exclusive breastfeeding for new-borns for at least, the first six months after birth (1,2). This is significant because it is the surest way to ensure good health and survival of infants. However, breast milk as biofluid is not pristine (3). Contaminants in breast milk can either be transferred from mother to foetus during intra-uterine development or through breastfeeding during lactation. Notable among environmental chemicals that contaminate human breast milk are polychlorinated biphenyls (PCBs) (4).

Polychlorinated biphenyls (PCBs) are mixtures of synthetic chlorinated hydrocarbons that have been used extensively since 1930 in a variety of industrial applications, such as dielectrics in transformers and large capacitors, as heat exchange fluids, as paint additives, in carbonless copy paper and in plastics (5,6). There are 209 possible congeners of PCBs. PCBs are among a group of 12 harmful organic chemicals known as the “Dirty Dozen” earmarked by the Stockholm Convention for elimination or severely restricting their production. A notably property of PCBs is general inertness. PCBs are resistant to acids and bases and possess thermal stability, making them useful in a wide range of applications, such as dielectric fluids in transformers and capacitors, heat transfer fluids and lubricants (7,8).

Chemical stability and lipophilicity of PCBs enable them to bioaccumulate and biomagnify in body tissues. In addition to their resistance to metabolism enhances their ability to persist in the environment for a long time. PCBs may associate with the organic components of soils, sediments and biological tissues, or with dissolved organic carbon in aquatic systems, rather than being in solution in water. Reports on human health effects of PCBs range from carcinogenicity, genotoxicity, reproductive toxicity, immunotoxicity and neurochemical effects (9). Research indicates that PCBs in humans can lead to increased rates melanomas, liver cancer, gall bladder cancer, brain cancer, gastrointestinal tract cancer, bacillary tract cancer and even breast cancer (10–16). Exposure to PCBs has the potential to supress the human body immune responses and thereby increase the risk of acquiring other human diseases (10,17,18). Association between elevated exposure to PCB mixtures and alterations in liver enzymes, hepatomegaly, and dermatological effects such as rashes and acne has been reported. Adverse effects are predominantly associated with higher blood concentrations.

Many people were exposed to PCBs and other toxins such as polychlorinated dibenzofurans (PCDFs) as a result of the contamination of rice oil by PCBs in Taiwan, China (1979) and Japan (1968). According to (19–21) pigmentation of the nail and mucous membrane is one of the signs and symptoms of exposure from these incidents, as well as enlargement and hypersecretion of the Meibomian glands of the eyes, swelling of the eyelids, and mucous membrane swelling. Hyperkeratosis and skin darkening with enlarged follicles and acne-like outbreaks ensued (frequently with staphylococcal infection) (22). Children born in the Taiwan event up to 7 years after maternal exposure showed hyperpigmentation, malformed nails, and natal teeth (23). Along with behavioural issues and increased levels of activity, they also had intrauterine growth retardation, worse cognitive development up to age 7, and intrauterine growth delay. At age 12, the impacted kids appeared to "catch up" with the controls. Children who were born 7 to 12 years after their mothers were exposed to PCBs had mildly delayed development, but their behaviour was not significantly different from that of children who had not been exposed (24). The problems seen in these kids are probably due to PCBs’ long-lasting effects in the body, which led to exposure during pregnancy years after the first exposure (24,25).

These effects are consistent with the observations of poorer short-term memory functioning in early childhood, in children exposed prenatally by mothers who had high consumption of Lake Michigan sports fish containing PCBs, amongst other persistent organic pollutants (POPs). People exposed in the Yucheng incident had low resistance to infections and suffered from a variety of them. Examination during the first year revealed decreased concentrations of IgM and IgA, decreased percentages of total T-cells, active T-cells and helper T-cells, but normal percentages of B-cells and suppressor T-cells; suppression of delayed type response to recalling antigens; enhancement of spontaneous proliferation of lymphocytes and an enhancement in lymphoproliferation to certain mitogens. After 3 years, some, although not all, of the effects had disappeared. Cancer deaths in both male and female workers involved in the manufacture of electrical capacitors were significantly increased. A significant increase in haematological neoplasms and gastrointestinal cancers was observed in male workers (9,19,20,26). Concentration factors in fish exposed to PCBs in their diet were lower than those for fish exposed to PCBs in water, suggesting that PCBs are bioconcentrated (taken up directly from the water) as opposed to being bioaccumulated (taken up by water and in food). The main source of PCB exposure to the general population is through food, especially fish (22).

PCBs concentrations in breast milk or in cord blood are dependent to the maternal PCB and dioxin body burden (27). This body burden is the result of PCB and dioxin-like accumulation over many years, especially in fat tissue, combined with low metabolic degradation and excretion rate (27). Most PCB monitoring studies were performed in Europe (28–32). Due to lack of standard measuring protocols, it is difficult to assess breast milk trends of PCB contaminants. However, some researchers have speculated that, over the last 25 years, levels may have decreased slightly (33). In Sweden, Czech Republic and Germany, where data have been collected following fairly consistent methods over time, evidence of a downward trend has emerged (28,31,32). Studies evaluating PCB levels in women found that breast milk concentrations were 4–10 times higher than in blood (33).

For example, in a study (34) reported a mean daily intake of 112–118 pg TEQ/kg bw in breast-fed infants and 6.3–6.5 pg TEQ/kg bw in 1- to 5-year-old children. “Maternal age is positively related, and the period of previous breast-feeding is negatively related to maternal PCB and dioxin-like concentrations” (27,35). Because more than 90% of the total daily human exposure to PCBs and dioxin-like chemicals is made up of oral intake from food, whereas other routes (e.g. water, air and soil) contribute less than 10% of total exposure (36), the role of diet has been investigated regarding breast milk pollution. In the U.S., fish consumption in the Great Lakes area has been associated with higher body burden of PCBs (37). In Canada, Inuit and fishing populations have higher PCB levels in breast milk than do urban population (Hirakawa et al. 1995). In the Czech Republic, the use of PCB-containing paint in grain silos resulted in higher breast milk PCB levels compared to neighboring regions and countries.

In a study (38) assessed the levels of PCBs in the breast milk of some Ghanaian women at suspected hotspot (Agbogbloshie) and relatively non-hotspot (Kwabenya) areas to ascertain if the levels of PCBs in mothers milk pose any health risk to the breastfed infants. A total of 128 individual human breast milk samples were taken from both primiparae and multiparae (Agbogbloshie – 105 and Kwabenya – 23) and the samples were analyzed using GC – MS/MS.

Mean concentrations of PCBs from Agbogbloshie (hot-spot area) and Kwabenya (non-hotspot areas) were 4.43 ng/g lipid wt. and 0.03 ng/g lipid wt. respectively. They found that PCB-28 contributed the highest of 29.5% of the total PCBs in the milk samples, and PCB-101 contributed the lowest of 1.74%. The estimated daily intake of PCBs and total PCBs concentrations in this work were found to be lower as compared to similar studies across the world. The estimated hazard quotient using Health Canada’s guidelines threshold limit of 1 μg/kg bw/day showed no potential health risk to babies. However, considering minimum tolerable value of 0.03 μg/kg bw/day defined by the Agency for Toxic Substances and Disease Registry (ATSDR), the values of some mothers were found to be at the threshold limit. This may indicate a potential health risk to their babies. Mothers with values at the threshold levels of the minimum tolerable limits are those who work or reside in and around the Agbogbloshie e-waste site.

Though several studies have determined PCBs in whole human milk, very few have examined the levels in the colostrum, transitional milk and mature milk simultaneously in the same study. To the best of our knowledge, no such study has been conducted in Ghana, West Africa, the whole of Africa and even in most part of the world. Apart from a Hungarian study (39) which determined PCBs and PCDDs in human milk during the three stages of lactation, studies of this kind are rare.

Breastfeeding mothers produce three different types of breast milk namely colostrum, transitional and mature milk during lactation. Based on these observations in the literature, we hypothesized that Ghana, which is rapidly becoming industrialized, motorized and urbanized, may be contaminated with high levels of OCPs. Therefore, we propose that these OCPs may bioaccumulate to different extents in the three types of breast milk produced by the nursing mother at levels exceeding the permissible safety levels set by the WHO.

Notwithstanding the fact that PCBs are essentially industrial-based chemicals and the Volta Region is non-industrialized, nevertheless, the global movement of PCBs is such that they can be transported everywhere. The study therefore, aimed to find out which portion of human breast milk would accumulate PCBs the most and to explore whether mothers delivering at the Ho Teaching Hospital had accumulated unusually high levels of PCBs in any of the types of breast milk which could pose significant health risks to their infants.

## Materials and Methods

### Study Design

The study employed a cross-sectional design approach to collect data. Determination of PCBs formed part of a broader study conducted at the Ho Teaching Hospital to quantify levels of contaminants in human breast milk. The study recruited a total of forty-seven (47) participants using purposive sampling.

### Study settings

The study was carried out at the Ho Teaching Hospital (HTH). The hospital was formerly known as the Volta Regional Hospital. The HTH was chosen as the study site because it has a wide public patronage. Besides its status as a teaching hospital, it also serves as a referral facility within the Volta Region. Below is the map of the study area (sampling site). **Figure 1** shows the sampling site.

**Figure 1:**
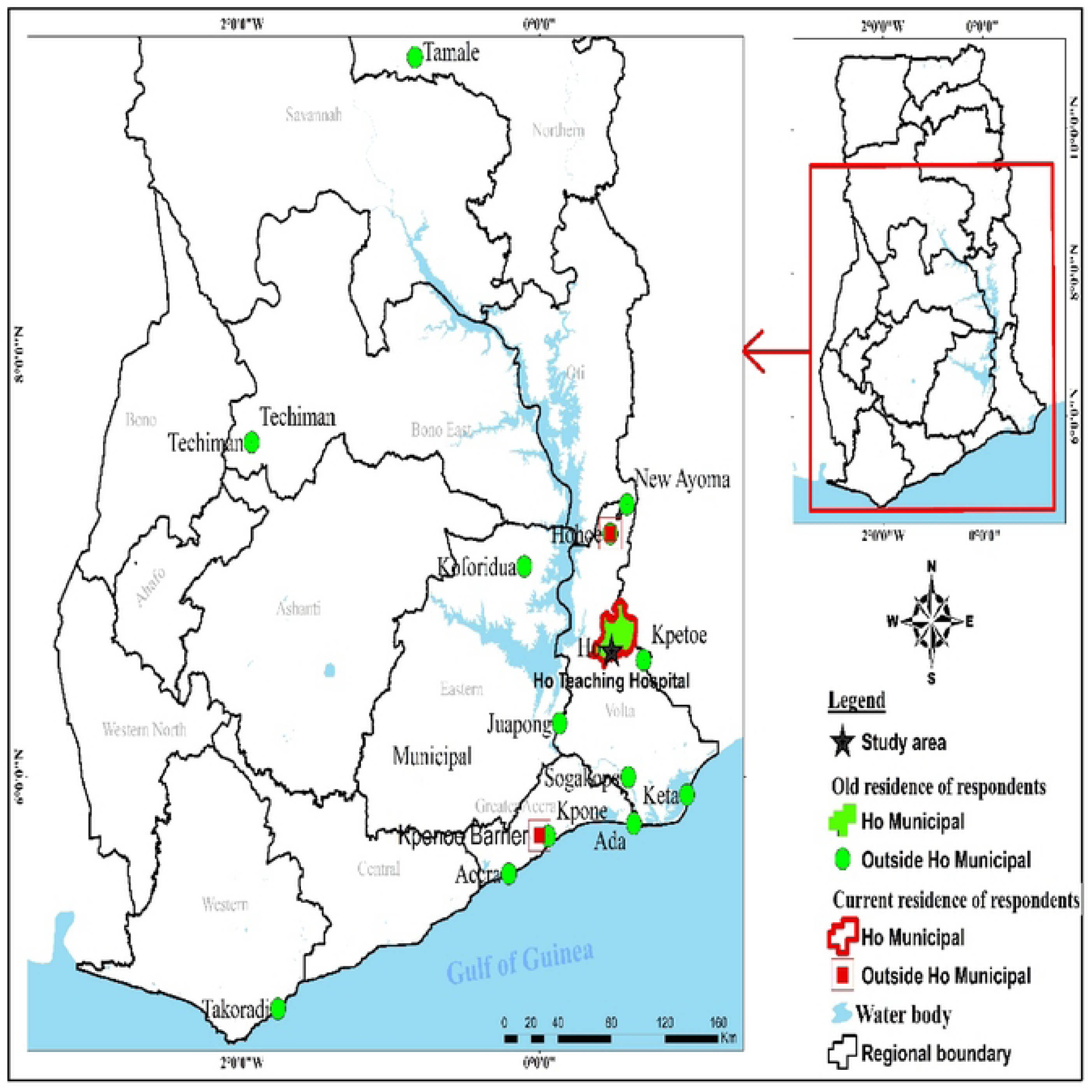
**A map of sampling site showing residences of participants**

Though most of the study participant were residing in the Ho Municipality during the time of the study, some of them had previously lived in other regions of Ghana ranging from 2 to 16 years.

### Study Population

The study recruited healthy lactating mothers aged 18 – 48 years who delivered at the Maternity Ward of the Ho Teaching Hospital during the time of the study; from December 25, 2020 to June 30, 2021. All healthy mothers in the age bracket 18 – 48 years who gave birth at the Maternity Ward of the Ho Teaching Hospital and were willing to take part in the study were included. Exclusion criteria included mothers who satisfied the above criteria but gave birth to twins and those mothers who could not be followed to their homes to collect the transitional and mature milks were excluded.

### Sample size and Sampling Method

The main sample investigated in the study was human breast milk. Though the estimated sample size was fifty (50) participants, the actual number of participants who willingly took part in the study was forty-seven (47/50); giving a success rate of sample collection of 94%.

Since we employed purposive sampling method, any mother who delivered at Maternity Ward during the time of the study and fell within the inclusion criteria and was willing to take part was selected. The process continued until we enrolled the 47 participants.

About fifteen millilitres each of colostrum, transitional milk and mature milk was collected from each participant during the different stages of lactation namely, from birth to day four (colostrum); from day five onwards up to 2 weeks (transitional milk) and from 2 weeks and beyond (mature milk). Arrangements were made with the midwife in charge of the Maternity Ward to facilitate collection of the samples. Colostrum milk was collected few hours after birth while the mother was in the hospital while transitional milk was collected during the one week postnatal discharge at the hospital. Thereafter, in cases where we did not get a participant to donate the mature milk at the facility, she was followed to her residence to do so. The process continued until we enrolled a total of forty-seven participants.

Besides the breast milk, each participant was made to complete a structured questionnaire to elicit information on biodata, geographical location and on dietary pattern. Since data were collected during the Covid -19 period, all Covid-19 prevention protocols were strictly adhered to during the sampling process.

### Ethical Considerations

The protocol for the study was reviewed and approved by the University of Health and Allied Sciences Research Ethics Committee (UHAS–REC), Protocol Number UHAS–REC A.9 [6] 19–20. Besides, permission and clearance were sought for from the relevant hospital authorities and departments before the commencement of the study.

### Materials and Reagents for PCBs Analysis

Mass Chromatograph (Agilent 7890B GC with Agilent Technologies GC Sampler 80), Mass Spectrometer (Agilent 7000C Triple Quadruple), Chromatographic grade ethyl acetate (pesticide grade, Merck), analytical grade acetonitrile, ACN, (99.8% from Sigma-Aldrich) sodium chloride (99.99%; BDH), anhydrous magnesium sulphate ACS (RG, 98.00%), trisodium citrate dehydrate (99.00%, Thermo Scientific^TM^), disodium hydrogen citrate sesquihydrate 99.00%, Thermo Scientific^TM^), primary secondary amine, PSA, (Thermo Scientific^TM^), formic acid, mixed PCBs standards obtained from Dr. Ehrenstorfer (GmbH, Augsburg, Germany). All the equipment including balances and pipettes were all calibrated to ensure accuracy.

### Sample Preparation for the Determination of PCBs from Breast Milk Samples

A modified version of QuEChERS was employed to extract PCBs from the breast milk samples. Approximately 10.00 g (± 0.05 g) of each sample of colostrum, transitional milk and mature milk was weighed into a 50.0 mL centrifuge tube, after which 10.0 mL acetonitrile (ACN) was added and then vortexed for one minute. Thereafter, a mixture of 4.00 g ± 0.20 g anhydrous magnesium sulphate, 1.00 g ± 0.05 g sodium chloride, 1.00 g ± 0.05 g trisodium citrate dihydrate and 0.50 g ± 0.03 g disodium hydrogen citrate sesquihydrate were added and vortexed for 1 minute and then centrifuged at 4000 rpm for 5 minutes. This was followed by dispersive solid phase extraction (DSPE). 6.0 mL aliquots of the ACN layer was then transferred into a 15.0 mL centrifuge which contained 150.0 mg primary secondary amine (PSA), 150.0 mg C18 and 900.0 mg MgSO_4_. The tube was closed and shaken vigorously for 30 seconds and centrifuged at 4000 rpm for 5 minutes. 4.0 mL of the cleaned extract was then transferred into a round bottomed flask and adjusted to about pH of 5 quickly by adding 40.0 μL of 5% formic acid solution in acetonitrile (v/v) and the filtrate concentrated below 40 ℃ on a rotary evaporator just to dryness. It was then re-dissolved in ethyl acetate by adding 1.0 mL using a pipette. The extract was transferred into a 2.0 mL standard opening vial for quantitation by a gas chromatograph (Agilent 7890B GC with Agilent Technologies GC Sampler 80 /MS (Agilent 7000 C Triple Quadruple). The extract was stored frozen until quantitation was achieved.

### Analytical Procedures for Determining PCBs from Human Breast Milk Samples

Analysis of PCBs from human breast milk samples was done using GC–MS/MS (Agilent 7890 B GC with Agilent Technologies GC Sampler 80 /MS (Agilent 7000 C Triple Quadruple).

## Quality Assurance Protocol

### Laboratory Reagent Blank

One laboratory reagent blank (made up of all chemicals except the sample) for each set of extracts were also run. The reagents were treated the same way as the samples, including exposure to all glassware and equipment that were used. The LRB were used to ascertain if method analytes or other interferences were present in the laboratory environment, the reagents or apparatus.

### Calibration Standards for PCB Congeners

Before analysing, the device was calibrated with a standard solution containing a combination of PCB congeners. The calibration standard solutions were purchased pre-packaged. Calibration was performed at seven (7) sites. Calibration standard solutions containing 5 ppb, 10 ppb, 20 ppb, 50 ppb, 100 ppb, 200 ppb, and 500 ppb mixed PCBs standards were used to calibrate the instrument in order to determine the detector’s linear response. The calibration point also include the LOQ. Table 1 summarises the GC - MS/MS operating settings for the study of PCB congeners.

**Table 1:** Summary of GC Operating Conditions for the Determination of PCBs.

| **Apparatus:** |  |
| --- | --- |
| **Instrument** | **Description** |
| Gas Chromatograph | Agilent 7890 B GC with a Agilent Technologies GC sampler 80 |
| Analytical column | 30m +10m EZ Guard x 0.25mm internal diameter fused silica capillary coated with VF-5ms (0.25µm film) from Agilent or equivalent. |
| Liner | Agilent 5190 -3167:900UL Single Tapered |
| Mass Spectrometer | Agilent 7000C Triple Quadrupole |
| **Temperatures:** |  |
| **Item** | **Conditions** |
| Injector | Splitless mode, temperature 280°C |
| Oven | 70°C / 2min <u>25°C/min</u> 150°C <u>3°C/min</u> 200°C <u>8°C/min</u><br>280°C/13.133min |
| MSD Transfer Line | 325°C |
| **Gases:** |  |
| **Gas** | **Flow rate** |
| Helium (carrier) | 2.25 ml/min constant flow |
| Collision gas<br>(Nitrogen) | 1.5 ml/min constant flow |
| Septum purge | 30mL/min at 0.75min |
| Pressure | 27.5psi |
| **Mass Spectrometry** |  |
| Segment start time | 4.0 min |
| Segment end time | 45.0 min |
| Source Temperature | 300°C |
| Scan Mode | MRM |
| Ionization mode | EI |

After establishing the GC operating parameters, the same settings were utilised to analyse the standards, quality control check samples, laboratory reagent blanks, and breast milk sample extracts.

### Performance Validation and Reproducibility

For the purpose of validating the method of extraction to identify matrix effects and determine the recovery of analytes or selectivity of the method, some samples were spiked with known concentrations of mixed PCBs standard and analysed to determine their recovery. The recovery studies of the PCB congeners ranged from 16% - 112%. Generally, higher PCB congeners gave very poor yields (PCB 209, PCB 194, PCB 180, PCB 170 and PCB 138 respectively) while the lower congeners like PCB 18, PCB 28, PCB 31, PCB 44 and PCB 153 gave higher yields for the recovery studies. Notwithstanding, the recoveries for the quality control samples were all good. **Table 2** summarises the recoveries of the spiked breast milk samples and quality control protocol standards.

**Table 2:** Performance of the GC-MS/MS Method for PCBs Analysis in Breast Milk.

| PCB<br>Congener | Retention<br>Time/min | LOD/<br>ppb | LOQ/<br>ppb | 50 ppb<br>Spiked | Recovery (%) of QCs |  |  |  |
| --- | --- | --- | --- | --- | --- | --- | --- | --- |
|  |  |  |  |  | 20 | 50 | 100 | 200 |

|  |  |  |  |  | ppb | ppb | ppb | ppb |
| --- | --- | --- | --- | --- | --- | --- | --- | --- |
| 18 | 11.853 | 0.0033 | 0.01 | 112 | 90 | 108 | 93 | 101.5 |
| 28 | 14.007 | 0.0033 | 0.01 | 96 | 95 | 100 | 106 | 94.5 |
| 31 | 19.885 | 0.0033 | 0.01 | 60 | 100 | 100 | 96 | 101.0 |
| 44 | 16.400 | 0.0033 | 0.01 | 84 | 75 | 100 | 96 | 109.0 |
| 52 | 15.495 | 0.0033 | 0.01 | 82 | 85 | 100 | 98 | 106.0 |
| 101 | 23.329 | 0.0033 | 0.01 | 80 | 85 | 102 | 104 | 99.0 |
| 118 | 23.329 | 0.0033 | 0.01 | 76 | 110 | 98 | 95 | 101.5 |
| 138 | 25.165 | 0.0033 | 0.01 | 56 | 95 | 102 | 94 | 102.0 |
| 149 | 23.937 | 0.0033 | 0.01 | 62 | 95 | 98 | 101 | 100.0 |
| 153 | 24.144 | 0.0033 | 0.01 | 58 | 90 | 102 | 96 | 101.0 |
| 170 | 27.420 | 0.0033 | 0.01 | 40 | 100 | 102 | 94 | 100.0 |
| 180 | 28.248 | 0.0033 | 0.01 | 32 | 80 | 100 | 95 | 106.5 |
| 194 | 29.979 | 0.0033 | 0.01 | 26 | 100 | 102 | 93 | 109.5 |
| 209 | 31.490 | 0.0033 | 0.01 | 16 | 75 | 102 | 94 | 101.5 |

### Calculation of Recoveries

Percentage recoveries of the spiked samples were calculated using the following formula:

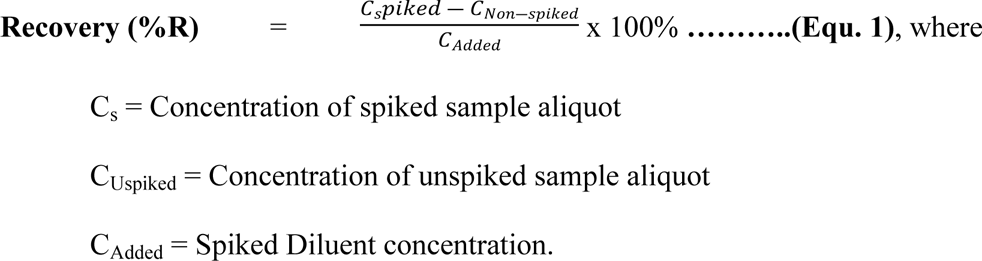

### The 14 PCB Congeners and their IUPAC names

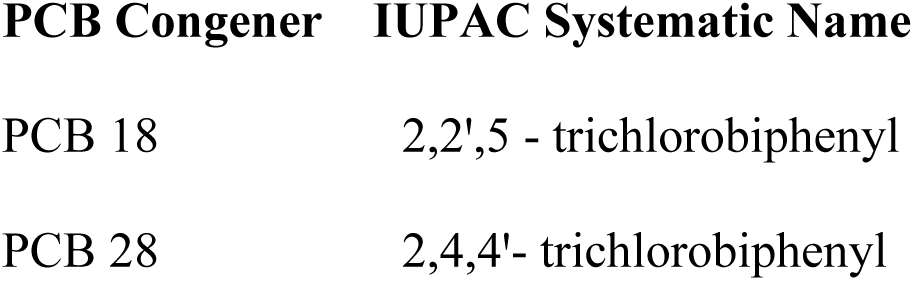
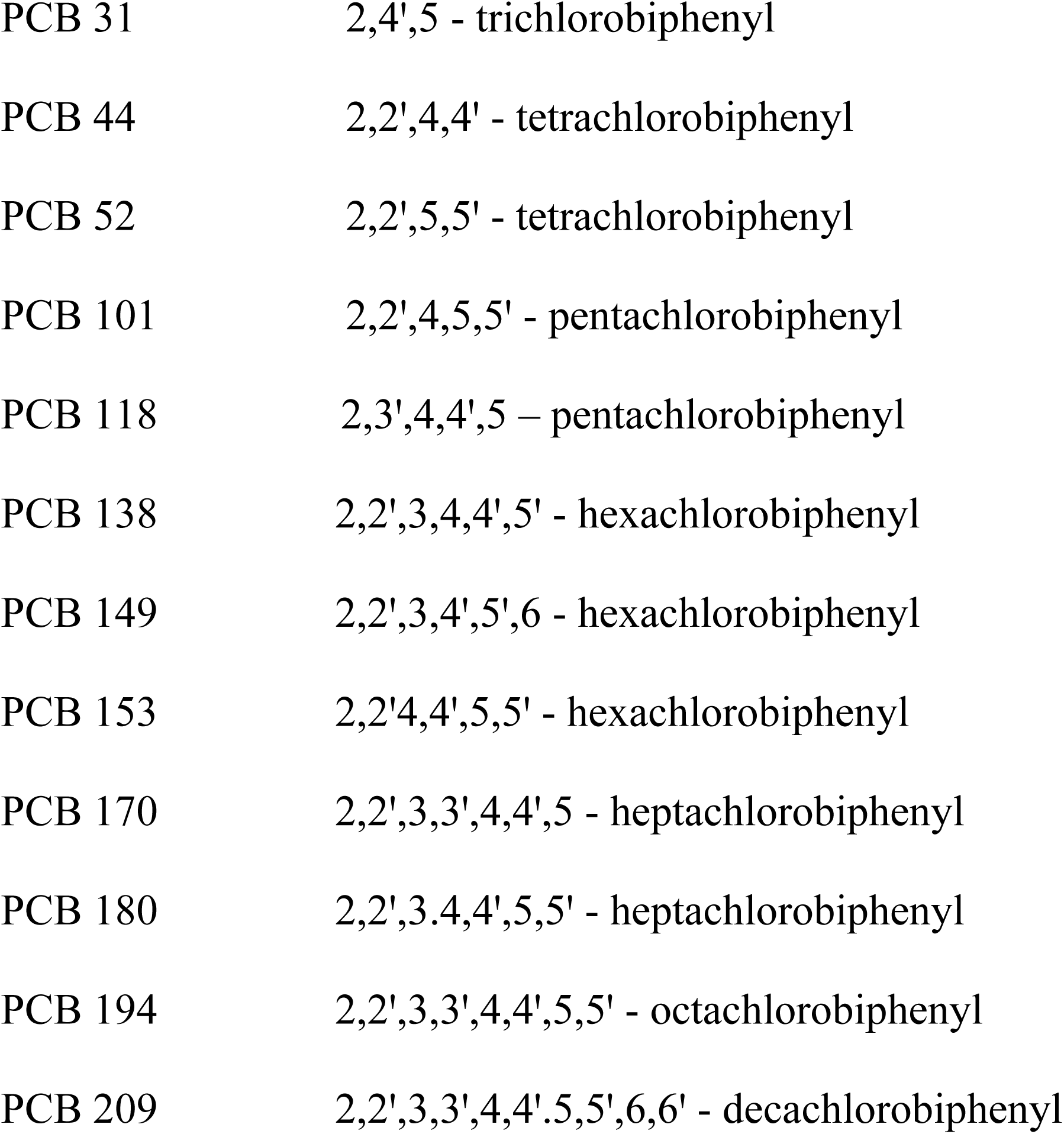

### Retention Time Windows

Retention time windows are critical for identifying target molecules. Absolute retention periods were used in addition to relative retention times to determine PCBs as congeners using the internal standard technique. To compensate for modest variations in absolute retention periods caused by sample loadings and typical chromatographic variability, retention time frames were created. The width of the retention time window was carefully chosen to reduce the possibility of both false positive and false negative outcomes.

### Quantitative Identification of PCB Congeners and Quantitation

The identification of PCBs as congeners using the GC - MS/MS method is achieved by matching mass to charge ratio between precursor and product ions of the known standards against the unknown sample and the use of retention time is also When a peak from a sample extract falls exactly within the retention time range established by a certain analyte of interest, this is referred to as tentative identification.

## Results

The results of the study are hereby presented and discussed. Forty-seven (47) fresh breastfeeding mothers were recruited for the study, representing 94% participation. The sociodemographic profile of the mothers is presented in Table 3.

**Table 3:**
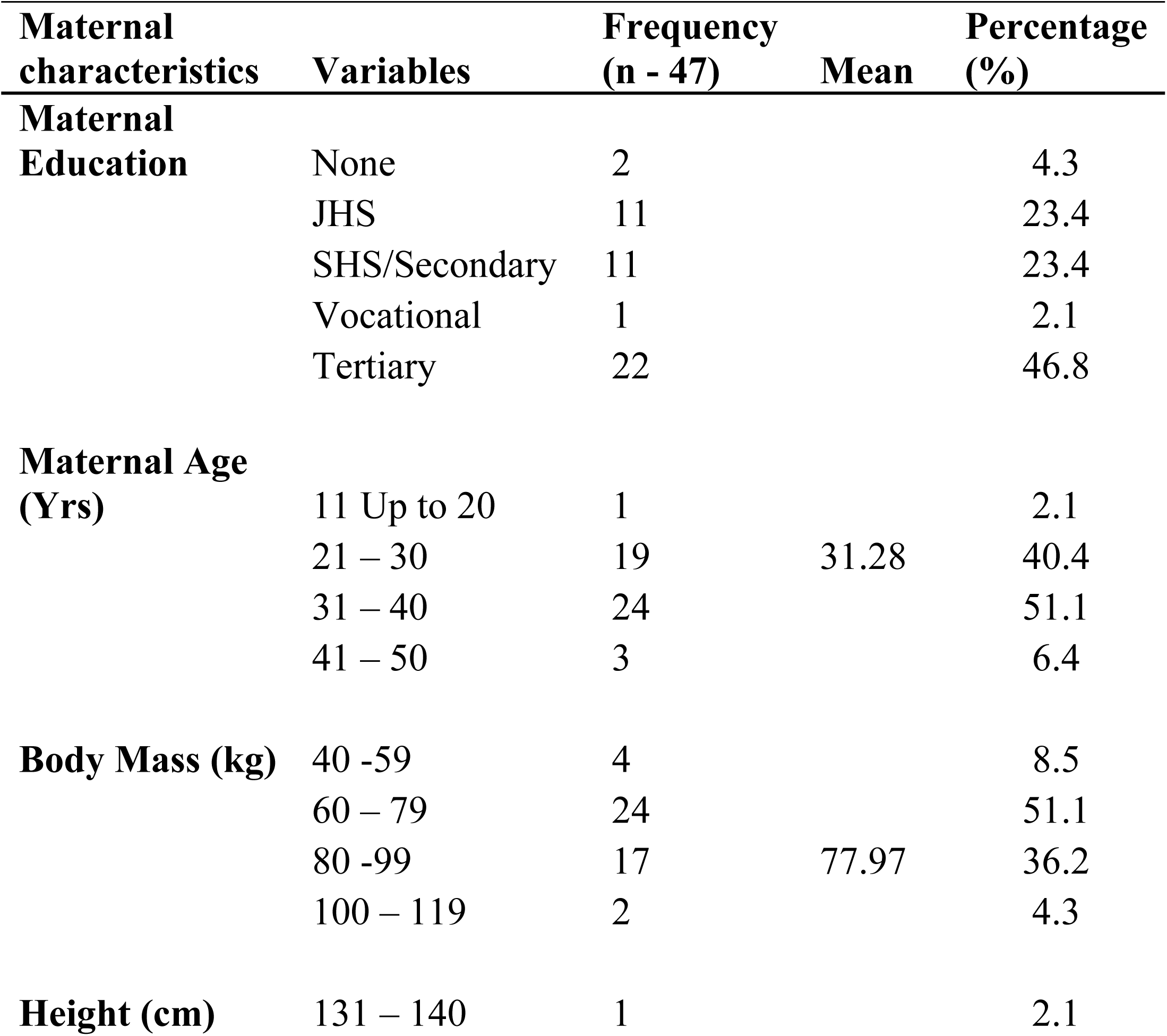
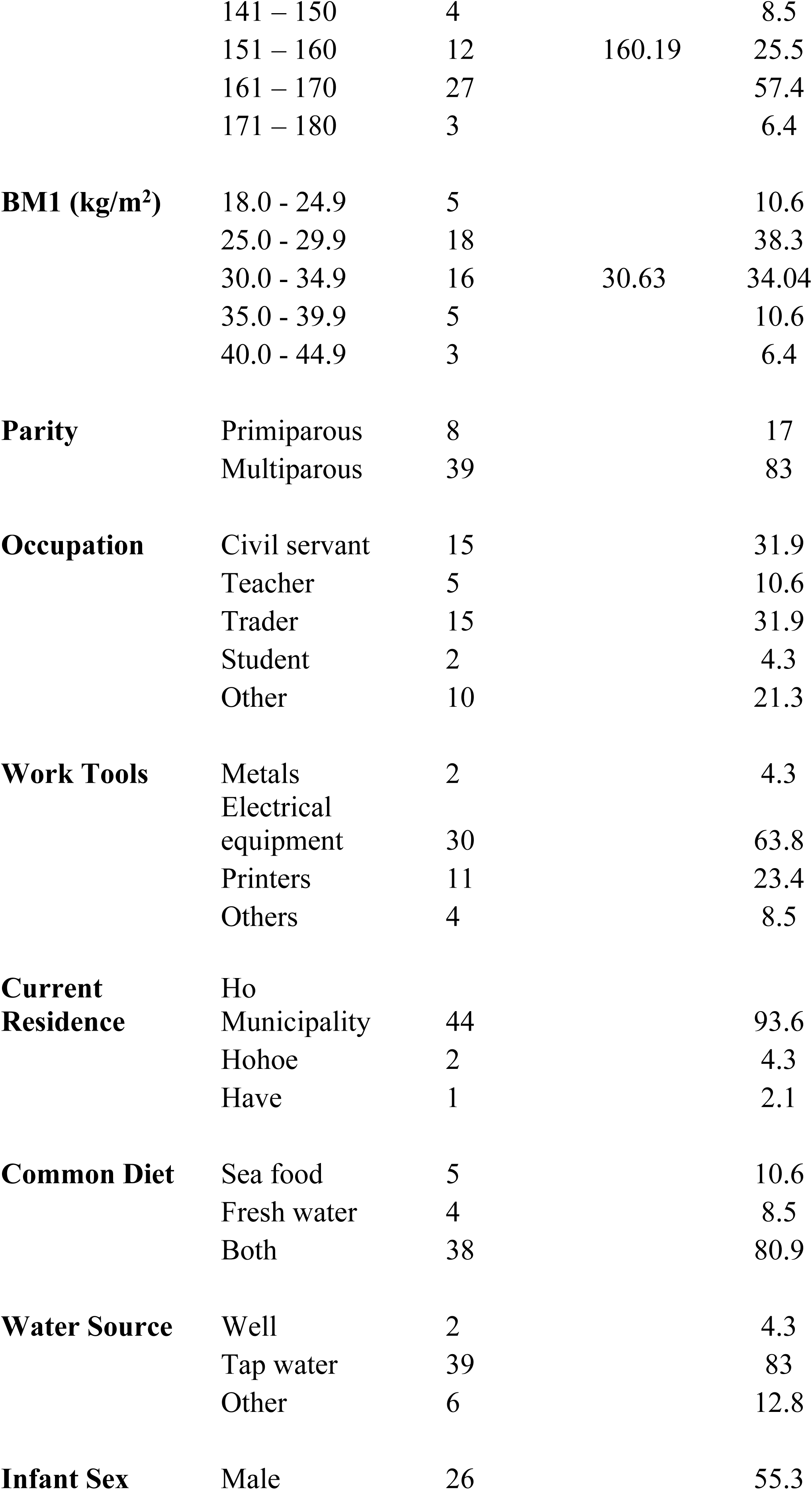
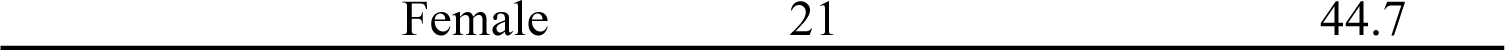
Sociodemographic Profile of Participants and their Babies.

The mean age of the mothers recruited for the study was 31.19 ± 5.48 years, with the minimum and maximum ages being 21 and 44 years respectively. Their average BMI was 30.63 ± 5.03 kgm_-2_. Of the forty-seven mothers recruited, 10.6% (5/47) had normal weight, 38.3% (18/47) were overweight while 51.0% (24/47) were obese. Most of the mothers were multiparous 39(83%) while the rest, 8(17%) were primiparous or first-time mothers.

### Levels of PCBs in Human Breast Milk

Forty-seven (47) triplicate breast milk samples made up of colostrum, transitional milk and mature milk from each participant, making up a total of 141 breast milk samples were analysed to determine the concentrations of 14 PCB congeners. Result of the analysis carried out on all the samples showed that all the PCB congeners were below limits of detection. The LODs and LOQs for the PCBs were 0.01 ppb and 0.03 ppb respectively. Since the PCBs levels in all in the portions of breast milk were below the threshold of detection, the background data of respondent were not subjected to statistical analysis.

## Discussion

The non-detection of PCBs in the breast milk samples analysed could be considered a positive development. The results of the study were probably an indication of the fact that the mothers in the study had low very low levels of exposure to PCBs. This could be explained by the fact that PCBs are essentially industrial-based chemicals and there are virtually no industries situated in Ho to cause the mothers to be exposed to these chemicals, even though PCBs as a member of POPs do not respect international boundaries (40,41). It could also mean that the campaign initiated by the Stockholm Convention on elimination of PCBs in the environment is yielding positive results. However, the non-detection should not be interpreted as a total absence of the PCBs in the mothers, and for that matter, in the environment. It may probably mean that some of the PCBs were present in the breast milk but their concentrations were not high enough to trigger or cause any significant health risks in the nursing infants. The ingestion of PCBs is of high toxicological relevance due to their long-term health effects. There is, therefore, the need for environmental samples to be analysed to understand the general pattern of distribution of PCBs in the area under consideration. While not dwelling much on their acute toxicity, research indicates that low levels of exposure to some PCBs may lead to possible endocrine disruption in humans (42), a situation which led to a ban on use of many PCBs for industrial and commercial purposes.

The study results were in line with a study conducted in Brazil (43) where they detected no contamination of human breast milk with PCBs in Rio Blanco/AC.

The study results were, however, in sharp contrast to those reported in Accra, Ghana by (38) who assessed the levels of PCBs in the breast milk of some Ghanaian women at suspected hotspot (Agbogbloshie) and relatively non-hotspot (Kwabenya) areas to ascertain if the levels of PCBs in mothers milk posed any health risk to the breastfed infants. A total of 128 individual human breast milk were sampled from both primiparae and multiparae (Agbogbloshie – 105 and Kwabenya – 23) and the samples were analyzed using GC – MS/MS. The total mean levels and range of Σ7PCBs were 3.64 ng/g lipid wt. and ˂LOD–29.20 ng/g lipid wt. respectively. Mean concentrations from Agbogbloshie (hot-spot area) and Kwabenya (non-hotspot areas) were 4.43 ng/g lipid wt. and 0.03 ng/g lipid wt. respectively. The present study site was, however, not regarded as a ‘hot-spot’ area for PCBs. Moreover, it is important to understand that the samples examined in the current were different from the ones employed in those studies.

It is important to recognize that the public health significance of PCBs contamination in human populations and the concomitant effects on breast-fed infants cannot be overemphasised. Some researchers have reported a decline in the levels of some POPs in human milk in recent times (44) which is a positive development in view of the negative effects of POPs on human population. Per the requirements of the Stockholm.Convention on POPs, levels of certain POPs in human breast milk would serve as an indication of the effectiveness of the treaty in eliminating or reducing emissions of selected POPs.

Even though the study results showed non detection of PCBs in human breast during three lactational stages, however, monitoring of PCBs in human breast milk is still important and should be encouraged. It is therefore, appropriate and relevant to inform and educate the general public about the harmful effects of such chemicals, and more importantly, the need of the government to ban or place restrictions on their use. This study will contribute significantly to efforts toward providing relevant data on current levels of PCBs in Ghana.

## Conclusion

The levels of polychlorinated biphenyls in the three portions of mothers’ milk examined at the Ho Teaching Hospital during lactation were all below detection limits. Thus the null hypothesis was upheld with regard to the distribution of PCBs in colostrum, transitional milk and mature.

Consequently, we concluded that the levels of PCBs in mothers’ milk were not high enough to pose any significant health risks to breastfeeding infants.

It is, however, recommended that environmental samples should be analysed to understand the distribution of PCBs in the area or region. Moreover, further studies should be carried out on human breast milk samples in other hospitals in the region to determine levels of PCBs during lactational stages.

Since prevention is always better than cure, there is the need for public education on the adverse health effects of PCBs to children and humans in general and how to protect the general population from being exposed to them in the environment that might lead to undesirable health consequences, especially, in breastfed infants..

## Authors’ Contributions

Conceptualization: JWAJ, DEKK, DKE, JKB, JT, HM; Investigation – JWAJ, DEKK, DKE, JKB, JT, HM; Funding acquisition – JWAJ, DKE, DEKK; Methodology – JWAJ, FOK, DEKK, JT & HM; Data analysis – JWAJ, DKE, JKB, DEKK; FOK; Resources – JWAJ; Writing and Editing – JWAJ, DKE, DEKK, JKB, FOK, HM; Project Administration - JWAJ

## Informed Consent Statement

Written informed consent was obtained from all participants who took part in the study.

## Data Availability

All data produced in the study are available upon request.

## Acknowledgement

We sincerely thank University of Health and Allied Sciences Research Fund and Ghana Standards Authority for funding this project. We are also grateful to the staff of Ho Teaching Hospital, especially, the Health Information Officer, Mr. Benjamin Amedume, and staff of the Ethics Department for their cooperation and support during the project. Finally, we wish to thank Ms. Norkplim Dei Hlorlewu and Mrs. Guide Mensah (midwives) sincerely for sacrificing so much in the collection of the breast milk samples, in spite of the challenges they faced.

## Supplementary materials captions

S1 Appendix: Questionnaire for potential breast milk donors

S2 Appendix: Informed consent form

